# Ex Vivo Human Liver Hyperperfusion Model: Study Protocol to Understand the Pathophysiology and Identify Strategies for Reduction of Small-for-Size Syndrome

**DOI:** 10.1101/2025.10.02.25337152

**Authors:** Shaili K Patel, Charalampos Konstantinou, Abdul R Hakeem, Tze Min Wah, Marina Karakantza, Richard Bell, Kin Cheung Ng, David J Beech, Laeticia Lichtenstein, Kondragunta R Prasad

**Affiliations:** Leeds Institute of Cardiovascular and Metabolic Medicine, School of Medicine, University of Leeds, LS2 9JT, UK; Department of Hepatobiliary and Transplant Surgery, Leeds Teaching Hospitals NHS Trust, Leeds, LS9 7TF, UK; Institute of Liver Studies, King’s College Hospital NHS Foundation Trust, London, SE5 9RS, UK; Department of Diagnostic and Interventional Radiology, Leeds Teaching Hospitals NHS Trust, Leeds, LS9 7TF, UK; Department of Haematology, Leeds Teaching Hospitals NHS Trust, Leeds, LS9 7TF, UK; School of Food Science and Nutrition, University of Leeds, Leeds, LS2 9JT, UK; Division of Transplant Surgery, Department of Surgery Medical College of Wisconsin, Milwaukee, WI 53226

**Keywords:** Hyperperfusion model, Small for Size Syndrome, Living Donor Liver Transplantation, Cancer Resection, PIEZO1, Liver Regeneration

## Abstract

**Introduction:** Liver has the unique ability to regenerate following surgical resection or partial liver transplantation. This underpins the surgical practice of cancer surgery and Living donor liver transplantation. A rate limiting factor for increased application of these techniques is the minimum volume of liver required for survival. Regeneration is triggered by increased portal blood flow and regenerating factors like hepatocyte growth factor through the remnant liver. However, an excessive blood flow, through a relatively smaller remnant liver has been postulated to cause injury to hepatocytes and failure to regenerate. This results in liver failure, a set of signs and symptoms together labelled as “small-for-size syndrome”. The underlying pathophysiology of injury and failure to regenerate is poorly understood. Most of the research is based on small animal studies, findings of which may not translate to human liver. The premise of this article is that a laboratory-based small-for-size graft model using human liver will enable a better understanding of the pathophysiology of increased blood flow, injury and regeneration. It is an opportunity to generate more directly translatable information.

**Methods and analysis:** Ex vivo human liver hyperperfusion model uses machine perfusion circuits to reproduce anatomical and physiological changes in livers that happen after a major resection or partial transplant. In this pilot study, whole liver normothermic machine perfusion is carried out for 4 hours followed by 6 hours of left lateral liver normothermic machine perfusion. This allows us to study effects and explore the role of altered mechanical forces (increased blood flow/pressure) on regeneration. Modulation of the proposed key mechanical force sensor (PIEZO1) is performed using agonist and antagonist drugs during this split liver perfusion.

**Ethics:** The protocol was approved by Health Research Authority and Health and Care Research Wales (23/NW/0361). It is endorsed by NHS Blood and Transplant, Research Operational Feasibility Group.

## INTRODUCTION

Liver disease is the eleventh leading cause of death (1) and liver cancer remains the fourth leading cause of cancer related mortality globally (2). The role of liver surgery, be it transplantation or resection is limited due to several reasons. These include, first, the shortage of available organs; second, the challenges of liver resection for cancer due to the small remnant liver; and finally, the complexities of Living Donor Liver Transplantation (LDLT) that are inherent from a need to balance donor safety with adequate remnant liver volume and recipient success with a minimum amount of functioning parenchyma. Addressing the shortage in organ availability is beyond the scope of our research. Novel technologies like machine perfusion are significantly advancing this field. The second and third limitations both involve the common factor of small remnant liver and its ability to function and regenerate, which is the main focus of our current research.

In the context of liver cancer resection, several strategies have been used to augment the remnant liver volume. These include portal vein embolization, associating liver partition and portal vein ligation for staged hepatectomy and dual vein embolization. While these techniques have helped in increasing the applicability of liver resection surgery in the management of liver cancers, they come with drawbacks such as increased waiting time, higher costs and need for additional procedures which contribute to morbidity and mortality. Importantly these approaches are not applicable in LDLT, where it is essential to preserve two functional portions with intact anatomical structure. To maintain adequate liver function, at least 30% of the total liver volume should generally be preserved. This threshold is calculated as the ratio of the future liver remnant (FLR) to the estimated total liver volume. Specifically, a standardised future liver remnant, which is the proportion of the FLR relative to the estimated total liver volume, of at least 20% is recommended after extended hepatic resections for patients with a healthy liver. However, for patients with conditions such as cirrhosis or hepatitis, a higher standardised future liver remnant of 40% is necessary due to the liver’s reduced regenerative capacity and baseline functionality. Simultaneously the recipient should receive a good volume of liver to ensure it meets the metabolic demands of their body. Currently the surgical decisions regarding safe remnant liver volume and minimum graft weight for successful LDLT are largely based on incremental modifications of the existing standards through trial and error. This has resulted in only modest expansions in the applicability of liver resection, whether for cancer surgery or for LDLT. This uncertainty stems from inadequate understanding of liver failure pathophysiology after surgery and from lack of research on human liver models.

Regeneration is triggered by increased blood flow through a small segment of the remnant or transplanted liver (3–7). Excessively high portal flow and pressure, termed as portal hyperperfusion, in relatively smaller liver segments can cause mechanical damage to cells lining liver blood vessels, also known as Liver Sinusoidal Endothelial Cells (LSECs), through excessive shear stress (8–11). Regeneration or tissue damage depends on the ability of small liver grafts or segments to cope with flow and pressure changes (12). Based on previous studies, living donor partial liver grafts with graft to recipient weight ratio <0.8% may not meet the metabolic needs of the patient and may lead to post-transplant hepatic dysfunction, also known as Small-for-Size syndrome (SFSS) (9, 13). The ILTS-iLDLT-LTSI consensus conference in 2023 defined SFSS as “a clinical syndrome caused by a partial liver graft that is too small to fulfill the metabolic demand of the recipient in the absence of specific surgical or nonsurgical causes” (14). The clinical syndrome is characterised by persistent cholestasis, increased ascites and coagulopathy (15).

Although the clinical effects are known, the regenerative mechanism underlying this imbalance remain poorly understood (16). Patients with SFSS have a higher mortality risk by up to 21%, and 34.7 to 40% reduced long-term survival compared to patients without SFSS, and this remains high in the absence of an optimal management strategy (17, 18). When looking closer at the microcirculation level, hepatic vascularization undergoes a series of important changes to allow flow adaptation and regeneration after hepatic resection. LSECs are activated by the increased blood flow following resection surgery (19). The consequent haemodynamic response is nitric oxide (NO) production to allow vasodilation as a result of mechanosensing from the fluid force (20). Recently discovered PIEZO1 proteins act as sensors of physiological mechanical force responding to flow in blood vessels (21, 22). Previous studies demonstrated the PIEZO1 protein’s role in the formation of new blood vessels and its presence in LSECs in mice and resected liver samples from patients (23, 24). Preliminary work on animal models showed that regeneration occurs at the molecular level by activation of PIEZO1 in LSECs and activation of Notch signaling (ADAM10/NOTCH1) mechano-sensitive pathway in mammalian cells (23, 25). Partial hepatectomy or associating liver partition and portal vein ligation for staged hepatectomy procedure in animal models have shown that shear stress induced endothelial nitric oxide synthase (eNOS[NOS3]) signaling plays an important role in liver regeneration cascade (7, 26, 27). Inhibition by NOS inhibitor N-nitro-arginine methyl ester reduced the short-term post operative proliferation factors, Ki-67 labeling index and future liver remnant/body weight ratio (7, 26). We have previously demonstrated PIEZO1 activation leads to NO mediated and endothelium dependent portal vein relaxation in response to mechanical and osmotic stretch (28). PIEZO1 agonist Yoda1 was used to specifically activate PIEZO1 independently of other potential force sensors and strong PIEZO1-dependent NOS3 phosphorylation was detected in isolated hepatic endothelial cells (29). Compounds such as Yoda1 offering the potential to modulate PIEZO1 proteins have recently been discovered (30–32). This has led us to hypothesize that PIEZO1 modulation may have a pivotal role in human liver regeneration after surgery.

Machine perfusion is increasingly being adopted as a technique to improve preservation, for viability testing of donor livers before transplantation and to recondition organs to improve short- and long-term outcomes (33–39). This technology can have extended applications for research (40).

Most of the research on SFSS is based on small animal studies, findings of which may not translate accurately to human livers. The premise of this article is that a laboratory-based human liver small-for-size graft (SFSG) model will facilitate studies that are more representative of the anatomical and physiological characteristics of the human livers, potentially leading to findings that are more directly applicable and translatable to clinical practice. This pilot study is designed to use machine perfusion technology to create a hypertensive state in a SFSG. Techniques, both surgical and ablative, will be used to reproduce anatomical changes to human livers on machine, as it would happen after a major resection surgery. We hypothesise that the same amount of blood flowing through a smaller segment (20-25% of whole liver) will simulate the excess perfusion that happens after extended resectional surgery. This will allow us to study the molecular mechanisms of SFSS in a human liver and initial pathways for regeneration, as well as uncover the mechanisms behind hyperperfusion injury and regeneration. Such understanding would be crucial to achieving the next advancement in liver resection surgery and partial graft liver transplantation.

## METHODS

### Study design

Ex vivo human liver hyperperfusion model to identify targeted therapies for prevention of SFSS is a pilot study to establish a laboratory based extended liver resection model to investigate portal hypertension after surgery. It is being conducted at a single site in Leeds through collaboration between the University of Leeds (UoL) and the Leeds Teaching Hospitals NHS Trust (LTHT). The protocol uses two stage Normothermic machine perfusion of human livers (NMP-L). The perfusion is undertaken at the UoL and the sample analysis at the UoL and LTHT.

### Organ procurement and consent

The deceased donor livers not suitable for clinical use following visual, histological or machine perfusion assessment are obtained through the National Health Service Blood and Transplant (NHSBT). The families are provided with appropriate verbal and written information by the specialist nurse-organ donation or donor coordinators. The use of liver for research is explained to the donor family, who are required to provide full and informed written consent to the NHSBT for the study to proceed. Non-clinical use group specific or O group leucocyte deplete packed red blood cells are obtained for perfusion from the local LTHT blood bank.

### Ethical and regulatory approval

Health Research Authority and Health and Care Research Wales approved this study (23/NW/0361). In addition, local hospital approval from Research and Development department at the LTHT and approval from NHSBT Research Operational Feasibility Group (ROFG) have been obtained. This study uses XVIVO Liver Assist™ device (Groningen, Netherlands) which has a Conformité Européene (CE) mark and is approved for non-clinical research use, Solero MWA device (AngioDynamics, Latham, NY, USA) and epoc® Blood Analysis System (Siemens Healthcare).

### Selection criteria

The deceased donor livers not suitable for transplantation are offered for research by the NHSBT. Suitable livers were selected for perfusion from September 2024 until the end of the study on or before 31 December 2026.

#### Sample size

12 whole human livers from deceased donors. This is a feasibility study and sample size estimation was based on number of livers offered for research by the NHSBT within the established timescale. As a feasibility study, the small sample size is not powered for statistical significance but will provide essential exploratory data to refine methodology and inform the design of larger, hypothesis-driven studies.

#### Inclusion criteria

- Adult donors (age 18-75), who have consented for research during organ donation
- Donation after Brain Death (DBD) or Donation after Circulatory Death (DCD: Cold ischemia time <16 hr for DBD, <10 hr for DCD and first warm ischemia time <60 min (from withdrawal) for DCD
- All livers with absence of Hepatitis B, Hepatitis C, and HIV

#### Exclusion criteria

- Age <18 years
- Pre-existing liver parenchymal disease like viral or autoimmune hepatitis
- Pre-existing extensive macroscopic liver steatosis, fibrosis or cirrhosis

### Procedure

Following selection and allocation of a suitable liver for research, the organ is transported in ice-cold preservation solution to the Leeds Institute of Cardiovascular and Metabolic Medicine (LICAMM), UoL by a medical transport for transplant services according to local protocol [Flowchart 1]. The research team requests units of compatible packed red cells from the blood bank for perfusion on machine. The liver is prepared and perfused with blood according to the established protocol [Appendix S1: Study Protocol].

**Flowchart 1:**
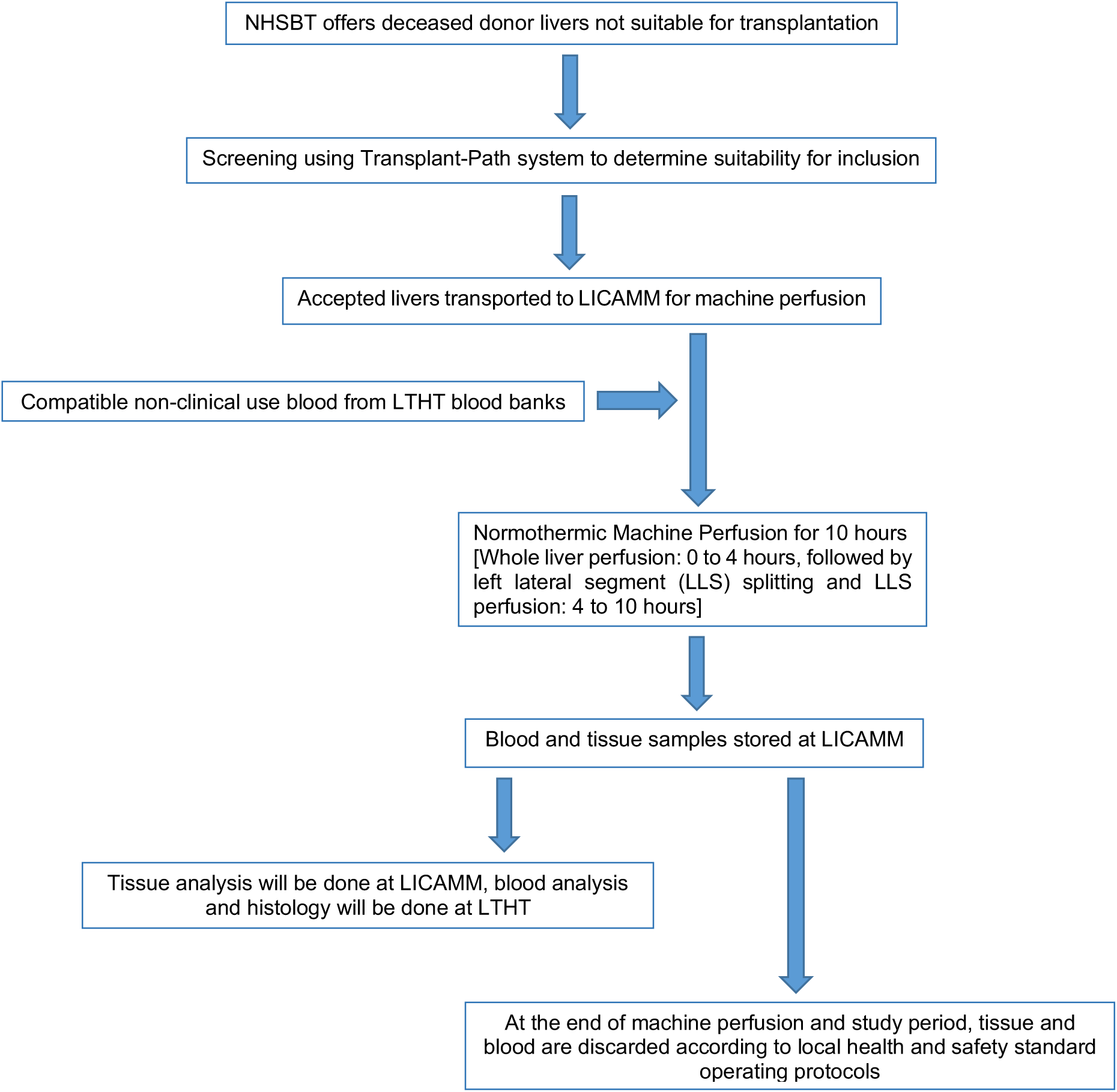
Process flowchart. From receiving offer to completion of machine perfusion. *NHSBT: National Health Service Blood and Transplant, LICAMM: Leeds Institute of Cardiovascular and Metabolic Medicine, LTHT: Leeds Teaching Hospitals NHS Trust*

#### Priming perfusion device

Machine is primed with perfusate suitable for normothermic machine perfusion (Table 1). The perfusion parameters are detailed in Table 2. Packed red blood cells and colloids are added to the machine via flow lines on the top of oxygenators. Antibiotics, Heparin, Sodium Bicarbonate and Calcium Gluconate are added to the perfusate. Parenteral nutrition, Epoprostenol, Insulin and Heparin are started as infusion.

**Table 1:** Perfusion fluid composition.

| Additives | Dose/Quantity | Volume |
| --- | --- | --- |
| Packed Red Blood Cells- Blood group specific or O group and leucocyte deplete | 3-5 units (hematocrit ~20-25%) | Approx. 1050 mL |
| Colloids (Succinylated Gelatin) | 2 units | Approx. 1000 mL |
| Antibiotics (Vancomycin & Gentamicin) | Vancomycin (500 mg) & Gentamicin (80 mg) | Approx. 10-22 mL |
| Modified Parenteral Nutrition (lipid free) | 4 to 15 mL/hr infusion | 150 mL |
| Fast acting Insulin | variable infusion | variable |
| Heparin | 10,000 IU followed by 1000 IU/hr infusion | Approx. 50 mL |
| Epoprostenol | 8 µg/hr infusion | Approx. 50 mL |
| 8.4% Sodium Bicarbonate | 30 mL followed by variable to balance pH | variable |
| 10% Calcium Gluconate | 10 mL | 10 mL |
| 50% Dextrose | 10 mL if perfusate glucose < 10 mmol/L | variable |

**Table 2:**
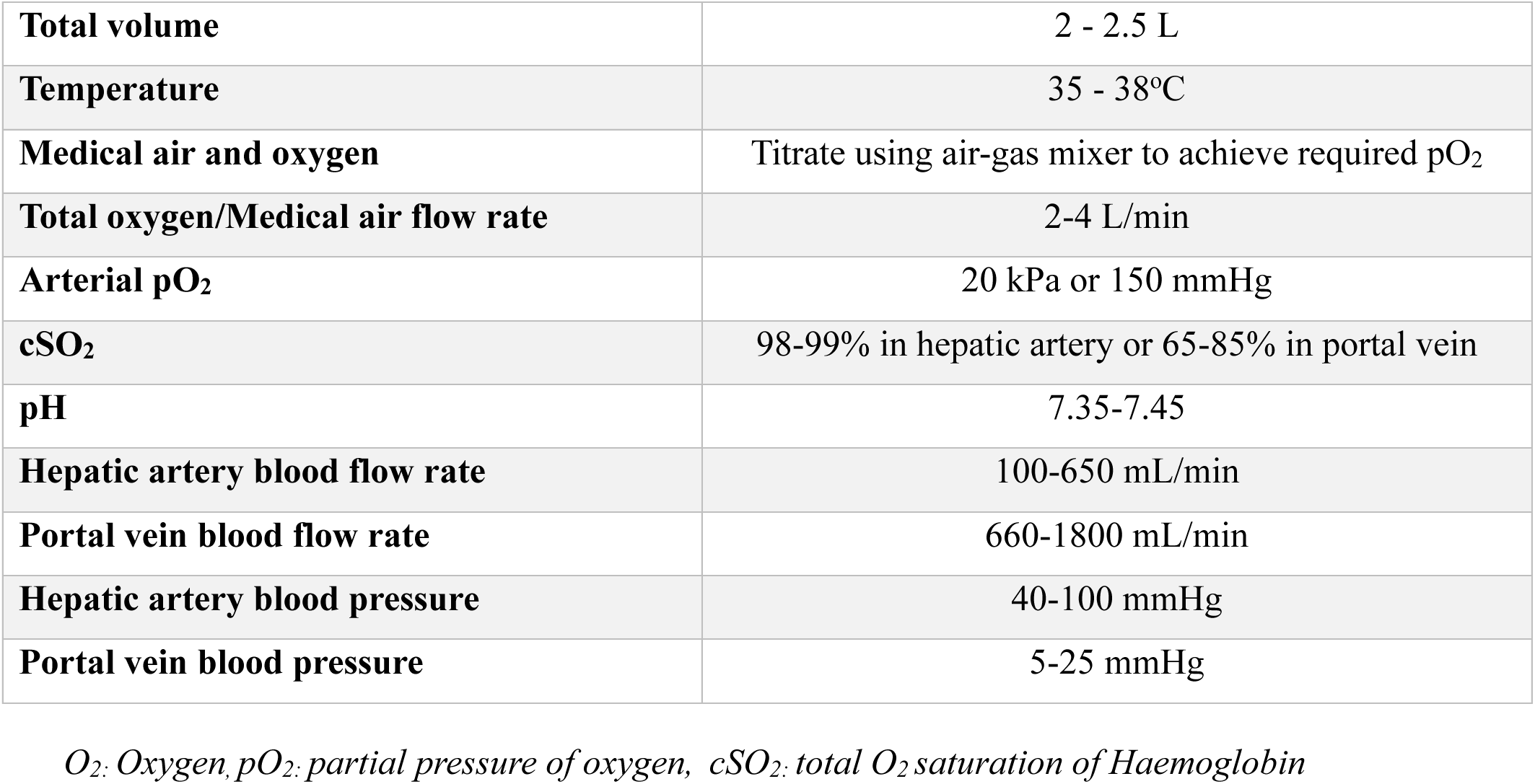
Perfusion parameters.

|  |  |
| --- | --- |
| **Total volume** | 2 - 2.5 L |
| **Temperature** | 35 - 38°C |
| **Medical air and oxygen** | Titrate using air-gas mixer to achieve required pO<sub>2</sub> |
| **Total oxygen/Medical air flow rate** | 2-4 L/min |
| **Arterial pO<sub>2</sub>** | 20 kPa or 150 mmHg |
| **cSO<sub>2</sub>** | 98-99% in hepatic artery or 65-85% in portal vein |
| **pH** | 7.35-7.45 |
| **Hepatic artery blood flow rate** | 100-650 mL/min |
| **Portal vein blood flow rate** | 660-1800 mL/min |
| **Hepatic artery blood pressure** | 40-100 mmHg |
| **Portal vein blood pressure** | 5-25 mmHg |

*O<sub>2</sub>: Oxygen, pO<sub>2</sub>: partial pressure of oxygen, cSO<sub>2</sub>: total O<sub>2</sub> saturation of Haemoglobin*

Tubing is cleared of all air bubbles, and venous and arterial pumps are turned on following manufacturer’s instructions. The pressure parameters are nulled against atmospheric pressure to ensure pressures measured during perfusion are real pressures in portal vein and hepatic artery and oxygenation is started.

#### Preparation of donor liver

The liver is prepared and weighed prior to placing on the machine. Hepatic artery and portal vein are dissected using scissors and side branches are ligated using surgical suture. Right hepatic artery is dissected from the bifurcation of hepatic artery proper and slinged with suture. Right portal vein is dissected at bi-/trifurcation and slinged beyond bifurcation.

Arterial and venous cannulas are inserted in the hepatic artery and portal vein respectively and secured with sutures. Hepatic veins remain uncannulated. A silicon catheter is inserted into the bile duct, secured with sutures and connected to a drainage bag.

Liver is flushed with 1000 mL of cold (0-4°C) and 500 mL of warm (37 °C) 0.9% sodium chloride. Liver is connected to the perfusion machine within 1-2 minutes of flushing.

#### Perfusion of liver

Total duration of the perfusion is 10 hours. Liver is positioned in the organ chamber with anterior surface facing down. To start the perfusion portal vein cannula is connected to the portal inflow tube of device and the hepatic artery cannula is connected to the arterial inflow tube of device. Arterial and venous flow, resistances and pressure rates are continuously monitored.

Whole liver is perfused for 4 hours (Phase 1). After 4 hours, ligation of the right liver vessels and ablation of segment 4 inflow is done to isolate left lateral segment (LLS; liver segments 2 and 3) while continuing the perfusion (Figure 1). Ablation probe for coagulation is placed 1-2 cm to the right of Falciform ligament to avoid any collateral damage to the left lateral segment. Cannulation of the main bile duct is continued to study bile output. Pressure and flow settings are adjusted to study the liver damage with high flow/pressure setting to simulate hyperperfusion. The parameters were chosen based on clinical and consensus guideline recommendations to reflect physiological relevant thresholds for SFSS risk. LLS perfusion is performed for 6 hours (Phase 2). During this second phase, vasoconstrictors, PIEZO1 agonist and antagonist drugs are infused to investigate effects on hyperperfusion injury.

**Figure 1:**
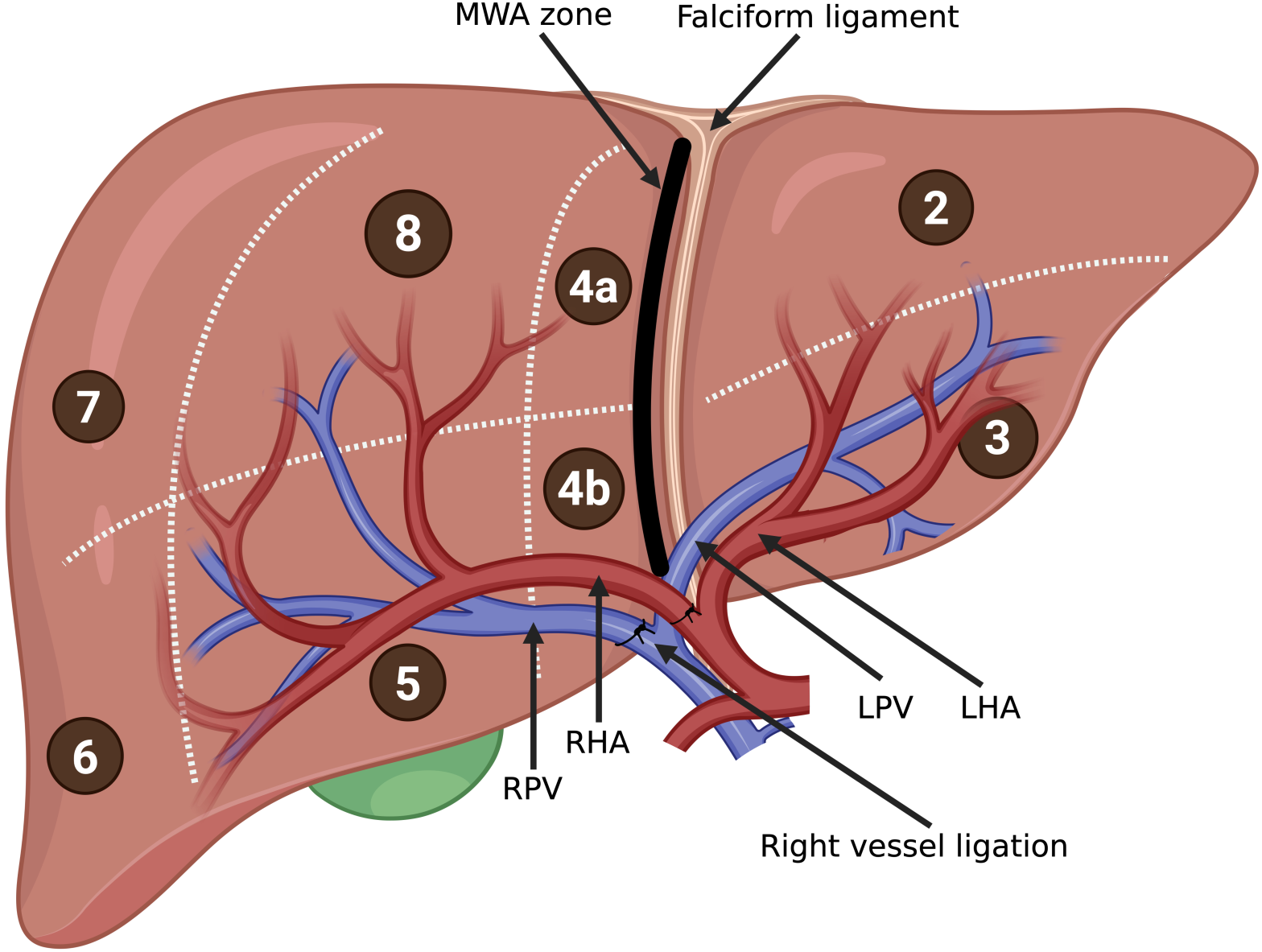
Left lateral segment (LLS) split. Segments 2 and 3 correspond to LLS. Right inflow vessels are ligated and ablation zone is created 1-2 cm to the right of Falciform ligament. *LPV: Left portal vein, LHA: Left hepatic artery, RPV: Right portal vein, RHA: Right hepatic artery, MWA: Microwave ablation.* Created in BioRender.

#### Collection and storage of samples

Tissue samples are collected, and blood gases, liver function tests and bile production are checked at the start of perfusion and at regular intervals (Table 3). Blood is collected in VACUETTE^®^ 3.5 mL Serum Gel red cap/yellow ring tube using sampling connectors and first 3 mL of perfusate is discarded as it is from the tubing. Serum is separated and stored in conical false bottom tube at - 80°C for biochemical analysis. Tissue samples are snap frozen in liquid nitrogen and store at -80°C for mRNA extraction and RNA sequence library preparation and Optimal Cutting Temperature or paraffin embedded for staining. Excision biopsy samples are stored in MACS^®^ Cell Storage Solution at 4°C for cell isolation.

**Table 3:**
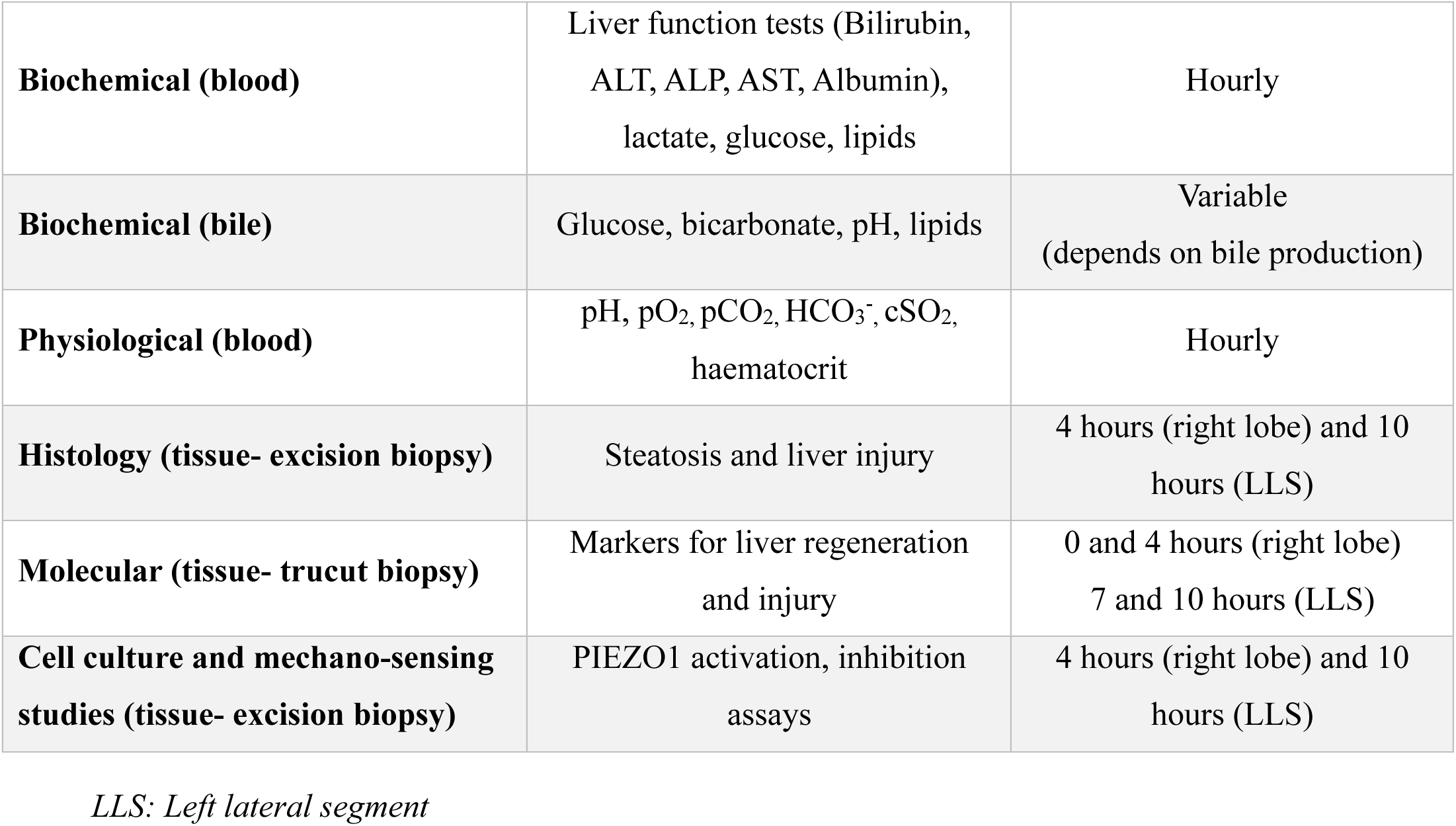
Blood and tissue analysis.

|  |  |  |
| --- | --- | --- |
| **Biochemical (blood)** | Liver function tests (Bilirubin, ALT, ALP, AST, Albumin), lactate, glucose, lipids | Hourly |
| **Biochemical (bile)** | Glucose, bicarbonate, pH, lipids | Variable<br>(depends on bile production) |
| **Physiological (blood)** | pH, pO<sub>2</sub>, pCO<sub>2</sub>, HCO<sub>3</sub><sup>-</sup>, cSO<sub>2</sub>, haematocrit | Hourly |
| **Histology (tissue- excision biopsy)** | Steatosis and liver injury | 4 hours (right lobe) and 10 hours (LLS) |
| **Molecular (tissue- trucut biopsy)** | Markers for liver regeneration and injury | 0 and 4 hours (right lobe)<br>7 and 10 hours (LLS) |
| **Cell culture and mechano-sensing studies (tissue- excision biopsy)** | PIEZO1 activation, inhibition assays | 4 hours (right lobe) and 10 hours (LLS) |

*LLS: Left lateral segment*

All samples are anonymized and stored securely at Leeds Institute of Cardiovascular and Metabolic Medicine, the University of Leeds in accordance with the Human Tissue Authority guidelines and local policies.

### Concomitant therapy

PIEZO1 agonist (Yoda1/Yoda2), PIEZO1 negative modulator (Dooku1) and Nitric Oxide Synthase inhibitor (L-NMMA) are added during the left lateral segment perfusion (Table 4).

**Table 4:** PIEZO1 modulating compounds.

| Compounds | Solvent | Maximum concentration | Dose Volume/2L perfusate |
| --- | --- | --- | --- |
| Yoda1 (Tocris) | DMSO | 10 μM | 2 mL of 10 mM |
| Yoda2 (Tocris) | DMSO | 10 μM | 2 mL of 10 mM |
| Dooku1 (Tocris) | DMSO | 10 μM | 2 mL of 10 mM |
| L-NMMA (Tocris) | DMSO | 20 μM | 10 mg |

*L-NMMA: L-N<sup>G</sup>-monomethyl-L-arginine, DMSO: Dimethyl sulfoxide*

### Objectives and outcomes

The primary objectives are:

1. Develop a laboratory based human liver hyperperfusion model using left lateral segment splitting technique.
2. To investigate the changes occurring in remnant liver from increased blood flow and pressure, and pharmacological modulation of mechanical regulation pathways.

Our secondary objectives are:

1. Biochemical profiling of perfused whole liver and hyperperfused segment
2. Histological characterization of perfused whole liver and hyperperfused segment
3. Transcriptomics, Lipidomics and PIEZO1 activity measurements for perfused whole liver and hyperperfused segment

### Analytical methods

Quantitative data is obtained on biochemical and metabolic functions of liver (Bilirubin, AST, ALT, ALP, Albumin, Lactate, Glucose), physiological function of blood and liver (pH, HCO_3_, pO_2,_ pCO_2_), bile production from liver and hepatic lipid homeostasis changes during perfusion. This data will be summarized as mean, median, standard deviation and ranges. Qualitative data is obtained on histology with Haematoxylin, Eosin, Oil Red O and Van Geison’s staining (evidence of steatosis, fibrosis and injury). Categorical data will be represented by numbers and proportions. Results for whole liver and split liver perfusion will be compared using t-tests or ANOVA.

Lipid profile will be tested using Liquid chromatography–mass spectrometry. RNA sequencing will be performed. We hypothesize gene expression changes for extracellular remodelling (e.g. HGF, Interleukin 6, TNFα, MMP 3-6), Serotonin and bile acid metabolism (e.g. CYP7A1, CYP8B1, CYP27A1), bile transporters (e.g. OATP1, MRP2, MDR3, NTCP), hepatocyte metabolism and growth (e.g. EGFR responsible for mitosis of hepatocyte, FGF involved in hepatocyte and stellate cells growth cycle), shear stress response (e.g. KLF2, ICAM1, PECAM1, eNOS), fibrosis response (e.g. COL1A1 and COL3A1), endoplasmic reticulum stress (e.g. ATF6, IRE1a and PERK), metabolic zonation (e.g. WNT, YAP, HYPPO), liver differentiation (e.g. HNF4a, FOXA1/2, SOX9, CK19), liver regeneration (e.g. VEGFR3, TGFα, PDGF, Beta1 Integrin, ESRP2, Interleukin 1), liver toxicity (e.g. GLP1, ARG1, GLS2, GST, CYP2C) and LSEC markers (e.g. Lyve1, CD31, STAB1/2, CLEC4G, FABP4/5). LSECs will be isolated and cultured in 96 well plates. Cultured LSECs will be stimulated with Yoda1 to measure PIEZO1 activity using Flexstation 3 (Molecular Devices) method where intracellular calcium concentration (i[Ca^2^+]) is measured.

### Risk assessment

#### i. Risk of handling human tissues and blood perfusate

Human tissue handling and blood perfusion carries a very small risk of transmission of blood-borne infections to the research team, estimated at less than 1 in a million units.

Only donor livers which are negative for Hepatitis B, Hepatitis C and HIV are accepted for this research study. The NHSBT routinely checks the donor virology for these transmissible organisms. The blood perfusate is obtained from local blood banks and has been extensively screened for clinical use, ensuring safely.

#### ii. Risk of machine technical failure

A CE-marked machine perfusion unit, which allows precise control of blood flow and pressure will be used according to the manufacturer’s guidelines. It has a small risk of malfunctioning or breakdown. As this is a laboratory-based project, there are no clinical risks to patients.

#### iii. Risk of leakage from cut-surface of liver

The liver may ooze blood from the cut surface when split surgically. Surgical splitting may increase the time required to meticulously control bleeding points. The use of ablative technology, minimises the risk of leakage, shortens the liver splitting time, and maintains an effective hyperperfusion model.

As there is no direct patient involvement, no additional risks to patients are anticipated.

### Data management

The consents are stored by the NHSBT in a safe and confidential manner. Confidential data is handled according to the Data Protection Act 1998 and guidelines for Good Clinical Practice.

Members of the Transplant Surgery clinical team have been authorized to access the clinical data through TransplantPath to ascertain whether the livers are suitable for inclusion in the study. They will identify suitable livers offered for research, pseudonymize them and allocate a unique Human Tissue Authority tracking reference number to each liver. The link to these study numbers is kept secure on the LTHT NHS password protected and encrypted computers. No patient identifiable information is stored on the LTHT or UoL premises. Publications will only use anonymized data. On completion of the study, the data will be stored in a secure archiving facility by the LTHT for 15 years.

### Study status

The study started recruitment on 3 September 2024. Recruitment, data collection and results analysis are expected to be completed by 31 December 2026.

## DISCUSSION

A study extracting data on primary liver cancers from 185 countries has predicted the number of new cases of liver cancer per year to increase by 55.0% between 2020 and 2040 (41). For primary liver cancers approximately 70% are diagnosed at a stage that prohibits conventional surgery or transplantation. This is often due to large disease burden precluding surgical resection (42). LDLT has an advantage over deceased donor liver transplantation in terms of shorter waiting times for recipient. Selection of appropriately sized donor graft is of prime importance to avoid SFSG related complications in both LDLT donor and recipient.

An important consideration in major resection or when using SFSG in LDLT is the prevention of SFSS. The vascular factors contributing to SFSS include recipient factors such as preoperative portal flow and pressure, and intraoperative factors such as portal flow (43) and pressure (44) , hepatic arterial flow (45) and hepatic venous outflow (46), with Portal Vein Pressure (PVP) being the most significant (8). Evidence suggests that PVP >20 mm Hg is detrimental (47, 48). Accordingly, portal inflow modulation is recommended in SFSG to achieve portal pressure <15 mm Hg and/or portal flow <250 mL/min/100g of liver tissue, thereby reducing the risk of SFSS, early graft loss, and poor outcomes^15^. Additional risk factors include graft quality (e.g., steatotic, fibrotic, or older donor livers), and recipient status (e.g., severe portal hypertension, acute liver failure, or acute-on-chronic liver failure). Metabolic dysfunction-associated steatotic liver disease (MASLD) is now the leading indication for liver transplantation in Western countries, and its high prevalence poses challenges for both donors and recipients. The degree of graft steatosis is also critical for preventing SFSS, although macrosteatosis up to 20% is generally considered acceptable for split liver transplantation (16, 49).

Research on animal models has shown overexpression of endothelin-1 and inducible nitric-oxide synthase and decreased gene expression for anti-oxidative stress proteins in SFSG (50, 51). These changes disturb the graft microcirculation, thereby increasing the PVP, inducing ischemia and making hepatocytes vulnerable to oxidative stress and exacerbate inflammation in the graft. The resulting effect may contribute to the pathogenesis of SFSS (52). It is also well known that bile acid signaling is essential for normal liver regeneration and in MASLD progression (53, 54). Our preliminary experience from animal studies has shown elevated PVP induced nitric oxide synthesis through activation of PIEZO1 channels (29). The same study also demonstrates PIEZO1 mediated lipid metabolism, upregulation of bile production and downregulation of hepatic cholesterol in PIEZO1 deleted mice. Based on these findings and the link between PIEZO1, portal flow and lipid metabolism, we hypothesize that PIEZO1 may be modulated to alter the post-surgical molecular responses affecting regenerative and lipid pathways in SFSG.

To our knowledge, this is the first protocol to investigate the pathophysiology of human liver regeneration in a hyperperfused liver segment using a machine perfusion model, while also exploring the potential role of protein targets in this process. The findings from this protocol are expected to improve understanding of mechanisms contributing to early liver failure after resection or partial liver transplantation, and to inform the development of therapies aimed at preventing SFSS, thereby improving patient survival. We encountered several challenges during study design, the foremost being recognition of the variability between DBD and DCD livers, with the latter grafts carrying a higher risk of primary graft dysfunction, and also poor regeneration potential. For the purposes of this study, it was also essential to include only livers without evidence of perfusion problems, either in situ or on back bench, as this could erroneously affect flow parameters studied.

The laboratory-based SFSG liver model offers several advantages but also has important limitations. While such models aim to replicate human physiology, they cannot fully capture the complexities of liver regeneration, immune responses, and vascular dynamics observed in vivo, which may limit the accuracy of extrapolating findings to clinical settings. Variability in donor factors such as age, graft quality, steatosis, perfusion status, and prolonged cold ischemia time are the common reasons for grafts being declined, all of which may further influence outcomes. The model may not account for such heterogeneity, potentially limiting the generalizability of study results. In addition, immune responses play a critical role in the development of SFSS during the intraoperative phase, but laboratory systems may struggle to effectively simulate these responses, particularly because the blood used in these models contains only packed red cells, lacking the full spectrum of immune cells and inflammatory mediators present in clinical transplantation. The limited perfusion time (around 6 hours) in laboratory models may not accurately replicate the longer-term behaviour of SFSGs in clinical settings. During this period, portal hypertension, a key factor in SFSG complications, may not be fully simulated, potentially reducing the relevance and translatability of the findings to actual liver transplant scenarios. Finally, accurately replicating portal flow, pressure modulation, and other vascular parameters remain constrained by current technological capabilities. Future refinements could include the use of leukocyte-rich perfusates to better mimic immune responses, extension of perfusion beyond 6 hours on the SFSG, and incorporation of pharmacological strategies targeting multiple mechanosensors beyond PIEZO1. Despite these limitations, this model remains the closest available platform for investigating the effects of liver hyperperfusion and offers valuable insights into the mechanisms underlying SFSS. By bridging the translational gap between animal experiments and clinical observations, this model has the potential to inform therapeutic strategies, optimise graft selection, and ultimately improve outcomes after major liver resection and living donor liver transplantation.

## Supporting information

Appendix S1: Study Protocol

## Supporting information

Appendix S1: Study protocol

## Data availability statement

Data supporting this study are included within the article and/or supporting materials.

## Authors’ contribution

Conceptualization: KRP, LL

Funding acquisition: SKP, DJB, LL

Investigation: SKP, CK, LL

Methodology: SKP, CK, MK, TMW, ARH, RB, KCN, LL, KRP

Project administration: SKP, CK, ARH, TMW, DJB, LL, KRP

Resources: TMW, DJB, LL

Supervision: ARH, KRP, DJB, LL

Writing– original draft preparation: SKP

Writing– review and editing: SKP, ARH, DJB, LL, KRP

