## Appendix S1: Study Protocol for "Ex Vivo Human Liver Hyperperfusion Model: Study Protocol to Understand the Pathophysiology and Identify Strategies for Reduction of Small-for-Size Syndrome"

**Affiliations**

**Supplementary Methods**

1. Priming the perfusion device 4
2. Preparation of donor liver 5
3. Machine perfusion of whole liver 6
4. Machine perfusion of hyperperfused segment 6
5. Tissue analysis 7

**Supplementary Tables**

1. Table S1: Equipment and Materials 3

#### Step-by-step protocol

### Table S1: Equipment and Materials

| Equipment and Materials | Company |
|---|---|
| Liver Assist | XVIVO, Netherlands |
| Liver Assist dual line perfusion set | XVIVO, Netherlands |
| Solero Microwave Tissue Ablation device | AngioDynamics, USA |
| Solero Microwave Tissue Ablation Applicator (14 cm) | AngioDynamics, USA |
| epoc Blood Analysis System | Siemens Healthcare, UK |
| EPOC BGEM BUN test card | Siemens Healthcare, UK |
| SuperCore Biopsy Instrument (14 ga x 15 cm) | Argon Medical Devices |
| One-piece Paediatric Arterial Cannula (12 Fr) | Medtronic |
| BD Primary Gravity Set with Stopcock | BD |
| VACUETTE® 3.5 mL Serum Gel red cap/yellow ring tube | Greiner Bio-One Ltd, UK |
| Syringes (Luer tip 5 mL, 10 mL) | BD Discardit |
| Blunt Fill Needle with Filter (18G x $1^1/_2$ inch) | Prosum |
| Prolene (3-0, 4-0, 5-0) | Ethicon |
| Mersilk (Sutupak 0) | Ethicon |
| Vicryl (Sutupak 3-0) | Ethicon |
| DeBakey dissecting forceps | - |
| Toothed dissecting forceps | - |
| Haemostatic forceps | - |
| Curved and straight dissecting scissors | - |
| Mayo scissors | - |
| Needle holder | - |
| **Medicines and Additives** | **Company** |
| Succinylated Gelatin (Gelofusin Ecobag 500 mL) | B. Braun |
| Vancomycin hydrochloride (1000 mg powder) | Demo S.A. |
| Gentamicin (80 mg/2 mL) | Wockhardt |
| Heparin sodium (10,000 units/10 mL) | Wockhardt |

| | |
|---|---|
| Sodium bicarbonate (8.4% w/v 100 mL) | B. Braun |
| Calcium gluconate monohydrate (10% w/v 10 mL) | Demo S.A. |
| Lipid-free parenteral nutrition (SmofKabiven Central, 900 kcal/8gN, do not mix lipid chamber) | Fresenius Kabi |
| Epoprostenol (Flolan 0.5 mg powder and solvent for solution for infusion) | GSK |
| Insulin (Actrapid 100 IU/mL) | Novo Nordisk |
| Sodium chloride (0.9% w/v 1000 mL) | Carelide |
| Sodium chloride (0.9% w/v 100 mL) | Fresenius Kabi |
| Dextrose (50% w/v 50 mL) | Hameln pharma ltd |

#### 1. Priming the perfusion device

1.1. Set the Liver Assist machine, connect the disposable dual line perfusion set and sensors using manufacturer's instructions.

1.2. Add 4 units packed red blood cells (~1100 mL) and Succinylated Gelatin (1000 mL) to the machine via the flow line on top of the oxygenators. Haematocrit should be >20%.

1.3. Power on and turn on the venous and arterial pump, following manufacturer's instructions.

1.4. Remove all air bubbles from tubing.

1.5. Null the pressure parameters against atmospheric pressure, this will ensure pressures measured during perfusion are real pressures in portal vein and hepatic artery.

1.6. Set the perfusion temperature at 37$^{o}$C, portal vein pressure at 7 mmHg and hepatic artery pressure at 70 mmHg. Once liver is connected set pressures to maintain total flow at 100-110 mL/100g/min for whole liver perfusion and at > 250 mL/100g/min for left lateral segment perfusion.

1.7. Start the oxygen at 0.5 L/min and medical air at 2 L/min.

1.8. Add Vancomycin (500 mg), Gentamicin (80 mg), Heparin (10,000 IU), 8.4% Sodium Bicarbonate (30 mL) and 10% Calcium Gluconate (10 mL) as bolus to the perfusate.

1.9. Reconstitute Insulin 2 mL (100 units/mL) in 100 mL 0.9% Sodium chloride to obtain concentration of 2 units/mL.

1.10. Reconstitute Heparin 10 000 IU in 40 mL 0.9% Sodium chloride to obtain concentration of 250 units/mL.

1.11. Take 8 mL of fully reconstituted Epoprostenol (0.5mg vial in 50 mL solvent, 10,000 ng/mL) and dilute in 42 mL 0.9% Sodium chloride to obtain concentration of ~2 μg/mL.

1.12. Start infusions for lipid-free parenteral nutrition (15 mL/hr), Epoprostenol (4 mL/hr or 8 μg/hr), Insulin (4 mL/hr or 8 units/hr) and Heparin (4 mL/hr or 1000 IU/hr).

1.13. Take perfusion sample for blood gas analysis 15 minutes after priming to check pH and electrolytes. Be sure to discard first 3 mL of perfusate as it will be from peripheral tubing. Titrate pH, blood glucose and oxygenation according to readings- this is done throughout the perfusion.

- If pH is <7.2 add 8.4% Sodium Bicarbonate (10 mL)
- If blood glucose is <10 mmol/L add 50% Dextrose (10 mL)
- If $cSO_2$ is <95% increase oxygen flow as required

#### 2. Preparation of donor liver

2.1. Dissect Inferior vena cava and leave both supra-hepatic and infra-hepatic ends are open.

2.2. Dissect the hepatic artery and portal vein using scissors and ligate the side branches using Vicryl surgical suture. Anastomose accessory or replaced hepatic arteries if present using Prolene to a suitable stump.

2.3. Dissect right hepatic artery from the bifurcation of hepatic artery proper and sling the right artery with Silk suture.

2.4. Dissect right portal vein at bi-/trifurcation and sling right portal vein beyond bifurcation with Silk suture.

2.5. Close Gall bladder defect with Prolene 3-0 and ligate cystic duct with Vicryl.

2.6. Record whole liver weight to set target flows.

2.7. Insert a 12 French arterial cannula (Medtronic) in the hepatic artery and secure with Silk sutures.

2.8. Insert a 24 French venous cannula (XVIVO) in the portal vein and secure with Silk sutures.

2.9. Hepatic vein remains uncannulated.

2.10. Insert a 12 Fr catheter into the bile duct, secure it with Silk sutures and connect it to a drainage bag.

2.11. Flush out the liver with 1000 mL of cold (0-4$^{o}$C) and 500 mL of warm (37$^{o}$C) crystalloid solution (0.9% Sodium chloride).

2.12. Connect the liver to the perfusion machine within 1-2 minutes of flushing.

#### 3. Machine Perfusion of whole liver (0-4 hours)

3.1. Position the liver in the organ chamber with anterior surface facing down.

3.2. Connect the portal vein cannula to the portal inflow tube of device and the hepatic artery cannula to the arterial inflow tube of device.

3.3. Start perfusion, setting parameters within range.

3.4. Take blood samples at the start of perfusion for baseline parameters and at regular intervals thereafter. Blood is collected using sampling connectors provided by manufacturer and first 3 mL of perfusate is discarded as it is from the tubing.

#### 4. Machine Perfusion of hyperperfused segment (4-10 hours)

4.1. After 4 hours, split into Left Lateral Segment (LLS- segment 2 and 3) while continuing the perfusion.

4.2. Ligate right hepatic artery. Continue cannulation of the main artery, now flowing into left hepatic artery.

4.3. Ligate right portal vein. Continue cannulation of main portal vein to perfuse left portal vein.

4.4. Continue cannulation of the main bile duct to study bile output.

4.5. Coagulate zone between left lateral and segment 4 of liver using microwave ablation device to seal blood vessels. Place ablation probe 1-2cm to the right of Falciform ligament to avoid any collateral damage to the left lateral segment.

4.6. Take blood samples at regular intervals till the end of study. Blood is collected using sampling connectors provided by manufacturer and first 3 mL of perfusate is discarded as it is from the tubing.

4.7. Pharmacological modulation is done by adding L-NMMA acetate, Yoda1/Yoda2 and Dooku1 to the perfusate at 8 hours 45 minutes from start of perfusion.

4.8. At the end of perfusion record liver weight for left lateral segment and remaining segments of liver.

#### 5. Tissue analysis

5.1. Take Trucut biopsies for RNA extraction and RNA sequence library preparation from right and left lobe at 0 hour as baseline samples, at 4 hours following normal pressure whole liver perfusion, at 7 hours halfway through left lateral segment hyperperfusion and at 10 hours at the end of left lateral segment hyperperfusion.

5.2. Take excision biopsies from right lobe for histology and cell isolation at end of 4 hours perfusion to reflect normal pressure whole liver perfusion.

5.3. Take excision biopsies from left lateral lobe for histology and cell isolation at end of 10 hours perfusion to reflect hyperperfusion in small graft.

5.4. Snap freeze tissue in liquid nitrogen and store at -80°C for mRNA extraction and RNA sequence library preparation or paraffin/OCT embed for staining.

5.5. Stain 10 µm sections of frozen livers or 4 µm paraffin embedded liver with Haematoxylin, Eosin, Oil Red O (lipid staining), and Van Geison for basic histology, lipid/collagen staining and perivascular fibrosis quantification.

5.6. Extract total RNA from individual samples and send for RNA sequencing or carry out real-time PCR. Measure eNOS, KLF2, ICAM1 and VCAM1 (indicators of shear stress response); Col1a1, Col3a1 (markers of fibrosis response), and CD31, CD34 (endothelial markers) and ATF6, IRE1a and PERK (endoplasmic reticulum stress sensors).

5.7. Isolate Hepatocytes and LSECs. Culture in 96 well plates to test PIEZO1 activity. Stimulate LSECs with Yoda1 at a concentration of 2uM and measure PIEZO1 activity Flexstation method where intracellular calcium concentration ($_i[Ca^{2+}]$) is measured.
